# Value sets and the problem of redundancy in value set repositories

**DOI:** 10.1101/2024.02.15.24302903

**Authors:** Sigfried Gold, Harold P. Lehmann, Lisa M. Schilling, Wayne G. Lutters

**Author notes:** Corresponding author. Email address. (SG).

## Abstract

**Objective:** Crafting high-quality value sets is time-consuming and requires a range of clinical, terminological, and informatics expertise. Despite widespread agreement on the importance of reusing value sets, value set repositories suffer from clutter and redundancy, greatly complicating efforts at reuse. When users encounter multiple value sets with the same name or ostensibly representing the same clinical condition, it can be difficult to choose amongst them or determine if any differences among them are due to error or intentional decision.

**Methods:** This paper offers a view of value set development and reuse based on a field study of researchers and informaticists. The results emerge from an analysis of relevant literature, reflective practice, and the field research data.

**Results:** Qualitative analysis of our study data, the relevant literature, and our own professional experience led us to three dichotomous concepts that frame an understanding of diverse practices and perspectives surrounding value set development:

1. Permissible values versus analytic value sets;
2. Prescriptive versus descriptive approaches to controlled medical vocabulary use; and
3. Semantic and empirical types of value set development and evaluation practices and the data they rely on.

This three-fold framework opens up the redundancy problem, explaining why multiple value sets may or may not be needed and advancing academic understanding of value set development.

**Conclusion:** The paper catalogues the methods and practices used and provides practical aid in managing the value set development process. It offers recommendations for improving that process and for software innovation in to support. In order for value set repositories to become more rather than less useful over time, software must channel user efforts into either improving existing value sets or making new ones only when absolutely necessary.

## 1. Introduction

Controlled medical vocabularies (e.g., ICD10, SNOMED, RxNorm, CPT, LOINC) catalogue clinical concepts and relationships between them. A concept is signified by an entry in a medical vocabulary generally consisting of a definition, one or more synonymous labels, and a *code* to identify the concept in representing specific clinical events in electronic health records (EHR), registries, claims databases, and clinical data warehouses. Value sets are groupings of these identifiers that facilitate data collection, representation, harmonization, and analysis. (We treat the term “value set” as more or less synonymous with “code set”, “concept set”, “code list”, and “enumeration”, which are also used in some contexts.)

This paper focuses on the use of value sets in the context of observational research using real-world data (RWD) [1]. Despite the use of hierarchical classifications and other data structures to signify concepts at different levels of granularity, value sets are almost always needed when querying clinical data sets since a phenomenon of interest can usually be indicated using a variety of different codes. A study algorithm to determine the relative likelihood of outcome O in patients experiencing condition C depending on their receiving treatment T_1_ or T_2_ will need to define cohort or phenotype algorithms for identifying patient records indicating O, C, T_1_, and T_2_. (Our use of the term electronic phenotype, or just phenotype, follows others in the field of observational research with RWD, e.g., [2–5], and can be confusing for those not accustomed to this usage. See Section 3.3 for a definition.) An essential step in such algorithms is to select patient records containing specified fields whose values are any of the codes in a value set. Though further temporal and conditional logic are often needed beyond the simple presence or absence of matching records in a patient’s digital chart, value sets are usually the starting point for phenotype or cohort algorithms.

Crafting high-quality value sets is time-consuming and requires a range of clinical, terminological, and informatics expertise. Scholarly and practical efforts to address challenges in value set management (i.e., helping RWD researchers identify and select the set of codes best fitted to their hypothesis testing and analysis goals) [3–12] have resulted in value set definition and documentation standards [13–17] and in methods and tools for authoring value sets [18–22], for assessing value set semantics and quality [23–29], and for enabling and promoting value set sharing and reuse [30–32]. These papers demonstrate problems of bias and inaccuracy in value sets shared on public repositories and many present specific methods to improve value set development. Williams, et al. [4]—in a paper we used as a seed article for our literature review—performs a comparative review of the value set literature, offering nomenclature, a consolidated articulation of published knowledge on value sets, and a valuable catalog of recommendations for advancing technology for managing value sets.

The current paper offers a view of value set development and reuse based on a field study of researchers and informaticists. We conducted an online survey, semi-structured interviews with a subset of survey participants, and observation where possible of participants working on value sets, finding a diversity in real-world value set development practices and perspectives previously unexplored in the literature.

While there seems to be universal agreement on the importance of reusing value sets (or phenotype definitions containing value sets), we have recognized through interviews and our own experience that repositories of these objects suffer from clutter and redundancy, greatly complicating efforts at reuse.

Value set repositories tend to contain many value sets with the same name or ostensibly representing the same clinical condition, making it difficult for potential re-users to choose amongst them. When multiple value sets are found, it can be difficult to tell if they are redundant, that is, if any differences among them are due to error or if there are principled reasons to define multiple value sets for certain phenomena.

It has been implicitly assumed that value set repositories would improve and grow in utility as they gained wider and longer use. We ourselves have claimed that repositories would benefit by cooperating to consolidate or centralize in order to generate positive network effects by attracting wider audiences [9]. As we demonstrate, the opposite appears to be the case. With ongoing use, repositories accumulate redundant and low-quality value sets, making it increasingly difficult for a potential re-user to identify high-quality value sets appropriate to their needs. Positive network effects will only accrue if all contributions to a repository are dedicated either to improving existing value sets or making new ones when absolutely necessary.

Qualitative analysis of our study data, the relevant literature, and our own professional experience led us to three dichotomous concepts that frame an understanding of diverse practices and perspectives surrounding value set development. These three dichotomies distinguish:

1. Permissible values versus analytic value sets. Permissible value sets are used in applications where data capture occurs (primary use.) Analytic value sets are used in analysis or research application (secondary use) in order to select records matching clinical conditions or events of interest.
2. Prescriptive versus descriptive perspectives on controlled medical vocabulary use. These tend to be held as implicit beliefs about coded concept, a prescriptive orientation is appropriate to permissible values contexts, while a descriptive orientation may be appropriate in secondary use, analytic contexts.
3. Semantic and empirical types of value set development and evaluation practices and the data they rely on. Semantic practices and data relate to vocabularies and meaning and are always necessary. A descriptive approach to identifying codes for an analytic value set, however, would require empirical analysis of patient-level data. Empirical analysis and validation are always desirable for analytic value sets, but it is frequently not feasible.

We will show how this three-fold framework opens up the redundancy problem, explaining why multiple value sets may or may not be needed (see 3.6.) Our field needs innovative software to help users navigate thickets of ostensibly redundant value sets not just to choose between them, but to make use of their differences in crafting value sets appropriate to researchers’ needs.

## 2. Methods

As noted, the intent of this research effort is to more deeply understand the diversity of real-world value set development practices, especially mapping the influence of specific contextual factors to those practices. The intended outcomes are both theoretical — developing a more precise, informative set of distinctions between approaches — and practical — providing guidance to informaticists to be deliberate in their decisions, thus enabling more accessible opportunities for both value set and process reuse in the RWD research community.

Our study design has been guided by the scholarly tradition of computer-supported cooperative work (CSCW). It is first predicated on the lived experience of the authors as reflective practitioners[33] with decades of experience creating and managing code-sets in research contexts. Their initial insight was bolstered or challenged through triangulation among three specific data collection activities: surveys, interviews, and participant observation.

Firstly, a custom, 21-question, web-based survey investigated participants’ experiences using value sets in the analysis of RWD. Recruitment focused on professionals with such experience, identifying them through the first author’s professional networks. Given the variety and inconsistency in nomenclature for RWD analysis elements and processes, questions were carefully balanced to capture differences in interpretation and use.

Secondly, a sub-set of survey participants were invited for a follow-on semi-structured interview. The purpose was to explore their value set authoring and reuse practices. The contextual nature of the interviews allowed them to demonstrate their tools and processes for developing value sets in person or via screen share.

The survey and interviews were approved by the University of Maryland IRB (#1405794-8). Recruiting began August 1, 2019 and ended on September 14, 2021. Taking the online Qualtrics survey required human subjects to read and sign our consent form. Interviewees signed a separate Qualtrics consent form. The survey and deidentified data are available at [34].

Thirdly, the first three authors acted as participant observers, embedded in key communities and numerous projects in this space, including OHDSI, PCORNet, Health Data Compass, the Army Pharmacovigilance Center, and the American Medical Informatics Association. While writing this paper, SG and HL worked on the National COVID Cohort Collaborative (N3C), observing and contributing to large-scale value set development and management efforts in a novel context. Their active participation in this wide range of projects has made them careful observers of value set development and curation practices.

The qualitative data collected from the surveys, interviews, and participant observations were content coded in NVivo through a process of analytic induction. Codes and emerging themes were iteratively developed with co-authors.

In this paper we present the unfolding interpretation of the results of this study as a dialogue among the literature, the reflective practice, and the field research data. The resultant theorizing yields a conceptual framework that is both *descriptive* (making sense and ordering the world as it is) and *prescriptive* (giving structure to practice to inform the world as it ought to be).

## 3. Results and discussion

### 3.1. Diversity of value set development contexts

Seventy survey invitations were sent out. Of the 49 responses, 36 were complete enough for analysis, yielding a response rate of 64% and completion rate of 47%. Table 1 shows the diversity of our sample population in terms of relevant demographic and work environment characteristics. Participants hold an array of degrees and work in a variety of disciplines. Most reported being involved in a small number of studies requiring value set development each year, working in teams of between 2 and 10 people, often from multiple organizations. Participants and their fellow team members brought a range of skills and expertise to these projects (Tables 1 and 2) and they worked on projects involving a range of vocabularies, domains, and data models (Table 3.)

**Table 1.** Participant demographics and work contexts; study and/or value set development roles played by participant and other team members.

| Participant Degrees | # | Studies conducted per year | # |
| --- | --- | --- | --- |
| PhD | 17 | 0 to 1 | 1 |
| MS/MA | 6 | 1 to 5 | 2 |
| MD | 5 | More than 5 | 1 |
| MPH | 2 | Team size (people) | # |
| BSN | 2 | 1 | 0 |
| RN | 1 | 2 to 5 | 2 |
| JD | 1 | 5 to 10 | 9 |
| Sector/industry | # | More than 10 | 1 |
| Academic | 12 | Team size (organizations) | # |
| Public | 9 | 1 | 1 |
| Academic professional | 8 | 2 to 5 | 2 |
| Pharma | 3 | More than 5 | 5 |
| Consulting | 4 |  |  |
| Discipline | # |  |  |
| Informatics | 22 |  |  |
| Clinical quality measurement | 8 |  |  |
| Economics | 3 |  |  |
| Software, epidemiology, ontology | 1 each |  |  |

| Study roles | Any team member | Participant alone | Participant with others | Others | No one |
| --- | --- | --- | --- | --- | --- |
| Analyst | 35 | 11 | 26 | 10 | 1 |
| Programmer | 35 | 10 | 20 | 15 | 1 |
| Statistician | 33 | 5 | 14 | 19 | 1 |
| Clinical expert | 31 | 3 | 5 | 26 | 3 |
| Informaticist | 30 | 11 | 22 | 7 | 4 |
| Investigator | 30 | 18 | 22 | 10 | 1 |
| Epidemiologist | 25 | 2 | 7 | 17 | 6 |
| Terminologist | 20 | 6 | 13 | 8 | 13 |

**Table 2.** Software tools, platforms, and repositories used in value set development and sharing.

| Software/tools used | # |
| --- | --- |
| R | 26 |
| SQL database | 24 |
| OHDSI/ATLAS | 17 |
| SAS | 13 |
| Python | 8 |
| Tableau | 7 |
| Epic | 6 |
| VSAC | 5 |
| i2b2 | 4 |
| Other | 13 |

**Table 3.** Vocabularies, vocabulary domains, and data models targeted by participants’ value sets.

| Vocabularies used | # | Data models used | # |
| --- | --- | --- | --- |
| ICD10CM | 29 | OMOP | 18 |
| CPT | 27 | PCORNet | 9 |
| ICD9CM | 26 | Local system | 9 |
| LOINC | 26 | Claim forms | 5 |
| SNOMED-CT | 25 | i2b2 | 4 |
| RxNorm | 24 | Other | 9 |
| HCPCS | 22 | Value set domains | # |
| NDC | 21 | Conditions | 34 |
| OHDSI/OMOP | 14 | Procedures | 30 |
| UMLS | 13 | Medications | 29 |
| MedDRA | 8 | Lab tests | 28 |
| PCORNet | 8 | Other | 9 |
| FDB | 5 |  |  |
| Other | 19 |  |  |

Green-shaded items are general programming or analysis tools, blue-shaded items are made particularly for working with medical data or value sets.

Of the nine most common tools our respondents reported using for value set development listed in Table 3, several are specifically designed for clinical or clinical research applications, providing support for authoring, sharing, and using value sets. The others are general programming and analysis tools with which value sets can be composed, evaluated, and used by linking to database resources containing vocabulary and patient information.

### 3.2. Diversity of value set development practices

Value sets and processes for developing them vary in many critical ways. The effectiveness of a given value set development process and the accuracy of the value set it produces depend as much on the thoroughness with which methods are applied as on the selection of those methods.

A particularly important factor shaping value set development practice is whether the value set is being developed for a single project, for use across multiple known projects, or for sharing and reuse in unknown future projects. Literature cited in the introduction [7,8,14,16–18,20,23–28] asserts the importance of reuse in addressing problems with value set quality. We suspected at the outset of this study that reuse was uncommon, as there is considerable evidence [9] that reuse is fraught with difficulties and that repositories accumulate many value sets ostensibly representing the same clinical phenomenon. Our field data (see Table 4) show 30 (83%) of our respondents reuse value sets made by others and 20 (55%) use repositories to find value sets for reuse. Many participants mentioned sharing value sets to public or private repositories — about a third to the Observational Health Data Sciences and Informatics (OHDSI) ATLAS web interface [35], a third to the Value Set Authority Center (VSAC) [31], and several to other repositories, publications, and research networks.

**Table 4.** Value set-related tasks performed by survey respondents or their team members.

| Value set-related tasks | # |
| --- | --- |
| Create value sets |  |
| Create value sets for local use | 28 |
| Create value sets for use by others | 27 |
| Vocabulary search | 15 |
| Vocabulary navigation | 16 |
| Consult clinical or terminology experts | 16 |
| Optimize value set to more parsimonious expression by replacing codes with intensional rules where possible. | 5 |
| Translate value sets across terminologies | 21 |
| Add value sets to repositories | 24 |
| Use existing value sets |  |
| Use value sets created by others | 30 |
| Use value sets from repositories | 20 |
| Use value sets from publications | 8 |
| Manual or automated comparison with existing value sets | 29 |
| Evaluation |  |
| Evaluate value sets (code-by-code or at the set level) | 26 |
| Approval for use by subject matter experts or other decision makers. | 15 |
| Review by terminology and clinical experts. | 18 |
| Empirical valuation |  |
| Examine frequently occurring codes in patients with phenomenon for possible inclusion. | 2 |
| Identify false negatives if a reference standard is available. | 7 |
| Identify false positives through chart review of matching patients. | 12 |
| If codes are semantically appropriate but absent in the intended data, they may be discarded as irrelevant or included for the benefit of future use. (Precalculated term usage counts sufficient.) | 2 |
| If prevalence of the target clinical condition is known, a significant discrepancy between prevalence and phenotype/value set result counts may be taken by developers to mean the value set requires further work before release. (Requires patient-level.) | 2 |
| Inspect patient and record counts. | 15 |
| Review patients matched by code(s) to confirm phenomenon. | 11 |
| Sanity check and review the count of patients matched by the whole value set — preferably by executing its containing phenotype algorithm. (Requires access to patient-level detail to generate value set counts.) | 13 |
| Sensitivity analysis of changes in results caused by modification of the value set. | 4 |
| Test value set on patient data, presumably in context of phenotype algorithm. | 13 |
| Other |  |
| If reference standard was used, report how it was made. | 2 |
| Report description and justification for validation methods used and any resulting statistics. | 3 |
| Informatics, standards, or infrastructure work related to value sets | 22 |

### 3.3. Permissible values versus analytic value sets

We distinguish two general types of value set based on the way they are used: *permissible value sets*, used for capturing clinical data in patient records, specifying code systems and code system values that can be entered into a particular data element. The items in a permissible value set might be presented to the user as a dropdown list or typeahead field, serving both to prompt the user with the allowable selection of values and prohibit entry of values not included in the set. *Analytic value sets*, on the other hand, are used in the analysis or querying of existing patient records to select those that are indicative of a clinical observation or event of interest where that phenomenon might have been captured using any of a number of codes. (In other contexts, such as data harmonization and clinical quality measures, value sets are used in more ambiguous ways that have both permissible and analytic qualities.) There are other use cases for value sets (see Table 5), but the differences between these two contexts (data capture and RWD analysis) will show why the distinction is needed.

**Table 5.** Contexts for value set development.

| Context | Reuse repositories | Permissible / analytic | Prescriptive / descriptive | Semantic / empirical evaluation |
| --- | --- | --- | --- | --- |
| Value sets for data capture | VSAC | Permissible | Prescriptive | Semantic |
| Other value sets for terminology services (FHIR, CTS2, etc.) | VSAC | Permissible | Prescriptive | Semantic |
| Clinical quality measures | VSAC | Both | Both | Mostly semantic |
| Single study, single database |  | Analytic | Descriptive | Both — but empirical is vital and possible |
| Network study or multiple related studies | ATLAS, N3C | Analytic | Descriptive | Both |
| For analytic reuse but not for a specific study, database, or question | N3C | Analytic | Descriptive | Mostly semantic |

The distinction is not about the digital structure of value sets or their definitions, but about the ways they are used and the practices appropriate to the development and validation of each type. While the distinction is not generally made in the literature or in value set repositories, our findings cannot be understood without drawing it. The HL7 definitions and other discussions of value sets tend to imply permissible as their archetypal use case [23,24,26,27,31,36–39,30,40–44]. While this paper covers both types, analytic are our primary focus [2–4,9,13,32,45–47], and a central claim we make is that analytic value sets necessitate different methods and tools to author, validate, share, and reuse value sets.

Fig 1 shows permissible value sets in context: a clinical data management system includes screens or forms, each of which will include data elements for capturing clinical phenomena like diagnoses, observations, and treatments. Data elements are defined in part by the values they are allowed to take. Specific screens and data elements in EHR, clinical trial, or registry applications may be focused on particular clinical phenomena such as diabetes complications or hypertension medications. A permissible value set then provides a list of subcategories or instances — e.g., cardiomyopathy or retinopathy, etc. for a diabetes complications data element — to populate dropdowns and constrain data element values.

**Fig 1.**
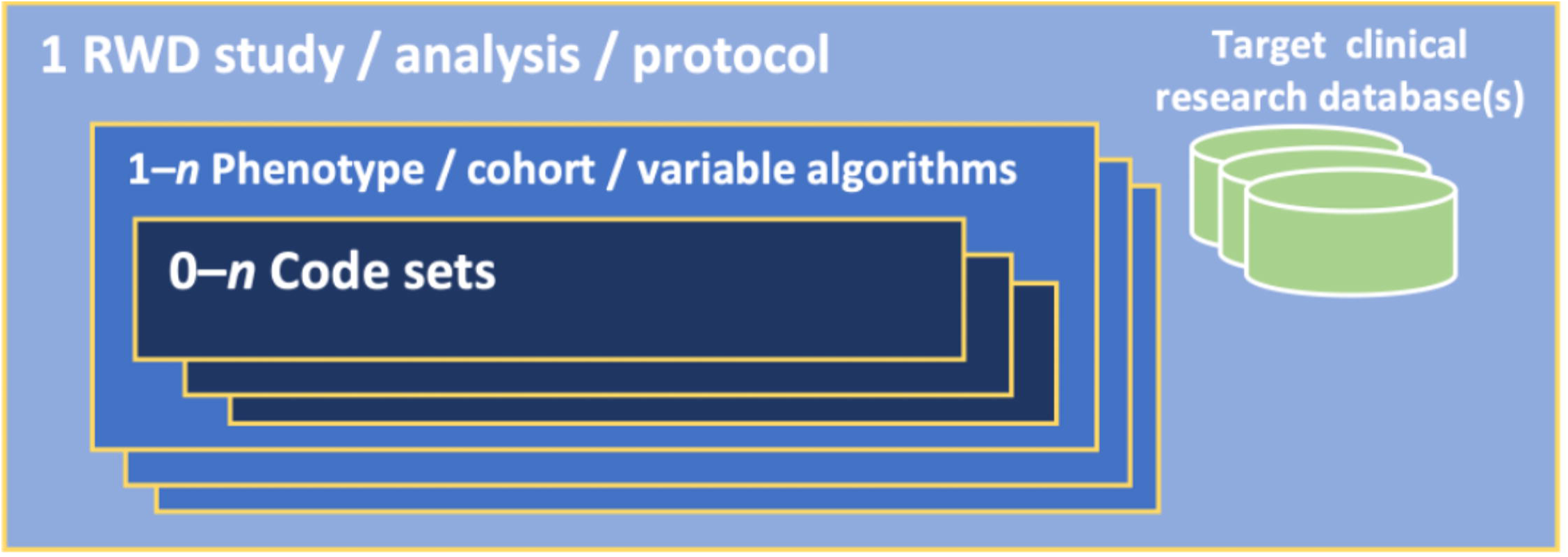
Permissible value sets in context.

In RWD studies, analytic value sets are used in the definition and identification of clinical phenomena of interest, representing study variables such as exposure and comparator cohorts, treatment or exposure criteria, process and outcome, covariates, confounders, etc.[3] The algorithmic components that identify specific clinical phenomena in the data may be called electronic phenotypes, phenotype algorithms, cohort definitions, or just variables; this paper mostly refers to them as “phenotypes.” Phenotype algorithms may use various types of data (narrative notes, images, EKG or other device output, etc.), but insofar as terminology codes are used in the algorithm, a phenotype will include one or more value sets as diagrammed in Fig 2 and may use temporal and conditional logic in performing set operations on the groups of patient records matched by different value sets. However, phenotypes can also be as simple as a single value set, the algorithm consisting of nothing but the selection of patient records containing one of the codes in that set. (See, e.g., “Finding Existing Phenotype Definitions” in the phenotyping chapter of the online textbook, Rethinking Clinical Trials [5] which lists value set repositories alongside repositories of more complex algorithms as sources of reusable electronic phenotypes.)

**Fig 2.**
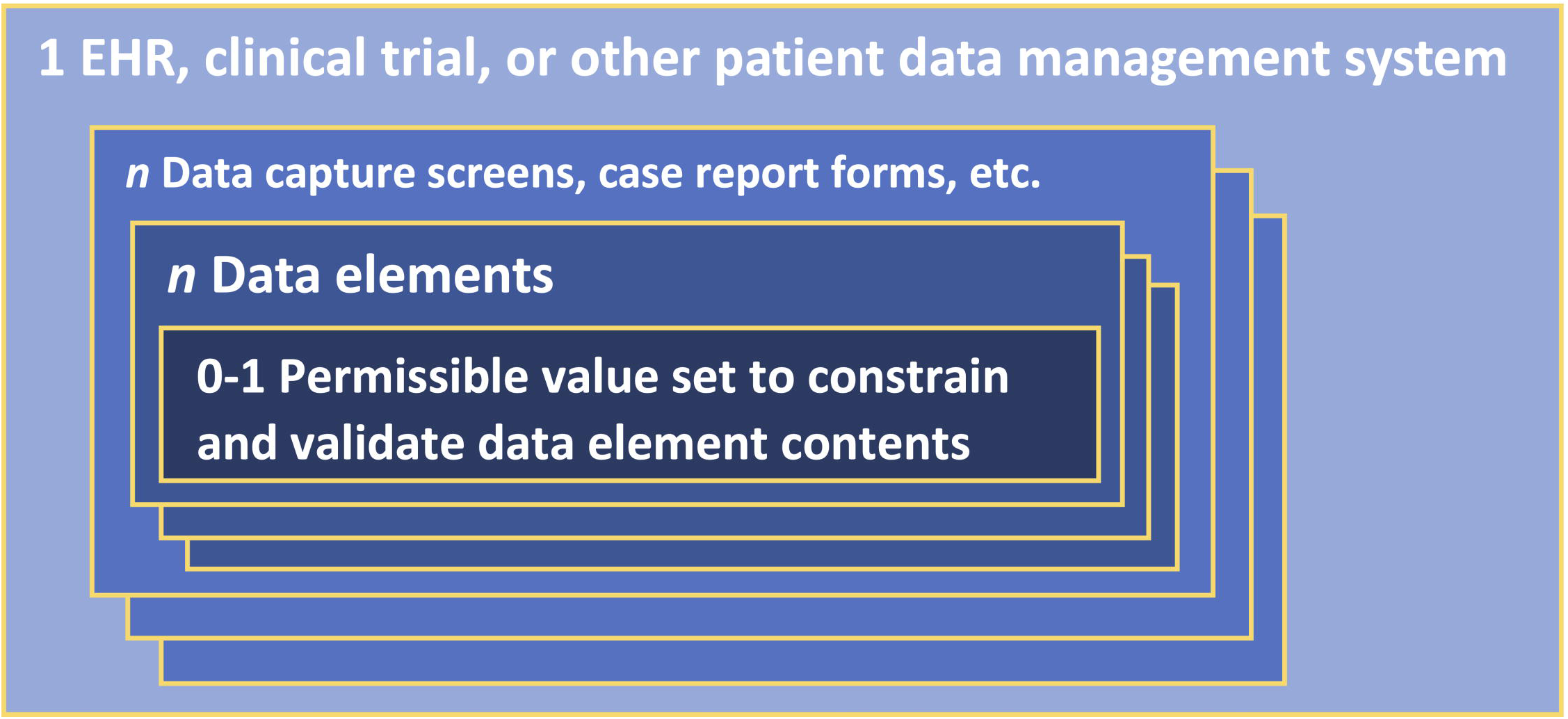
Analytic value sets in context.

Permissible value sets are generally developed for specific clinical data systems, often for a single institution. Analytic value sets are usually designed to reflect nuances of a particular research question, such as a need for sensitivity or specificity or the need for study-specific exclusion criteria.

### 3.4. Prescriptive and descriptive perspectives on value sets

Distilling and analyzing the catalogue of value set development practices in Table 4 — how value sets are made, used, evaluated, and reused — led us to re-evaluate the literature around value sets, observing that it falls into three research focus areas, which often do not seem to be in dialog with each other:

1. On permissible value sets for permissible values or clinical quality measures (CQM) [23–27,30,31,44,36–38,40–43,39]; Although value sets are not used as permissible value constraints in CQMs, the value set literature does not treat them any differently and does not address issues around aligning them with patient data — perhaps because CQM use by health care organizations is often required by payers, so alignment of value set and patient data is the responsibility of provider organizations, not value set developers.)
2. On phenotypes (cohort selection) for RWD research [2,3,13,46,47]; and
3. On analytic value sets (often called code sets) for use in phenotyping applications [4,9,19,32,45].

The computable phenotype literature sometimes conflates phenotypes and analytic value sets. Where it does discuss reuse, the focus is on phenotype rather than value set reuse. While some in this group do not believe that sharing value sets separately from phenotypes is worthwhile, we have seen that reuse does occur and there is demand for value sets that can be used across phenotypes. The permissible value set and value set literatures both tend to focus on value set repositories and reuse.

The permissible value set literature is not concerned with adapting value sets to specific clinical databases and looks to expert review or published sources of authority for value set validation. The phenotype literature, on the other hand, evaluates value set correctness primarily through empirical analysis with clinical data. The analytic value set literature falls somewhere in between. (The current paper shares more in common with this third group than the others, but it draws on all three. Those who identify, explicitly or implicitly, with one of these groups will benefit from being also informed by the others.)

Whether one considers expert authority or empirical analysis as the primary means of value set evaluation can reflect almost ideological beliefs about the nature of value sets and medical vocabulary use generally. Outside the domain of medicine and controlled medical vocabularies, in lexicographical or grammatical terms, the dichotomy between *prescriptivist* views and *descriptivist* views is well-known. For prescriptivists, dictionary entries and grammatical rules define how words and language *should be* used; proper language should conform with such rules and definitions. For descriptivists, dictionaries and grammars are attempts to capture a snapshot of how words and language *are* used in a given time and milieu. Non-conformant usage patterns indicate that the rules are lacking, not that the usage is wrong.

While terminological prescriptivism in natural language is generally considered unscientific and pedantic [48], the imposition of prescriptive terminology is, of course, the foundational purpose of standardized medical vocabularies and arguably the foundational practice of medical informatics generally [49–52].

Exemplifying a prescriptivist orientation, Winnenburg, et al. 2013 suggests that a value set should be anchored in a single concept, consisting of that concept and its descendants. That view is rejected by the descriptivist perspective held by many RWD researchers, expressed in rather extreme terms by one survey respondent:

> Code sets are always context specific. There is no such thing as diabetes in an RWD data source, there might be 50 definitions of diabetes and you have to pick the one that matches your question, data, and methods…We may spend months developing a code set for a specific question, iterating on different algorithms until the investigator is satisfied that the definition matches the needs of the study. (P04)

The following section distinguishes semantic from empirical techniques in value set development and validation. We have observed in a minority of our participants and in some of the literature a bias towards either semantic or empirical techniques that seems to override consideration of context and to reflect an implicit commitment to prescriptivist or descriptivist perspectives on controlled vocabulary use. That is, a person holding one of these views can find it difficult to see merit in the alternative. Overall, prescriptivist views should be appropriate for permissible **value set**s and data capture contexts; and descriptivist for analytic value sets and RWD research.

### 3.5. Semantic versus empirical methods and resources

Williams 2017 alludes to a central tension in the choice of methods for value set validation [4]. On the one hand, a rigorous validation would be to compare patient selection results against a reference (“gold”) standard created through medical record abstraction (MRA.) On the other, creating such a standard can be prohibitively time-consuming and require data that may be challenging or impossible to obtain. In our data, evaluation fell into two major categories, which we label overall as semantic and empirical.

#### 3.5.1. Semantic methods and evaluation by authority

We asked participants, “How do you verify that you have selected the best codes for representing a clinical concept in your analyses?” and received a range of answers. For many, confidence in their code selection came by reusing existing value sets from “previous[ly] published results” (P01), “examin[ing] the literature for validation studies” (P41), or “validated codesets when possible” (P04). (Value sets are available from value set repositories such as VSAC, ClinicalCodes [32], or the OHDSI/ATLAS or N3C concept set editors [35,53,54]); published papers that follow RECORD and other data-based observational study reporting guidelines [13,15]; previous projects available to the value set developer; and groupers such as Clinical Classifications Software Refined (CCSR) [55,56].

Other participants described an evaluation or validation process based on review by terminologists and clinicians (P19), clinical experts (P26), or “our coding panel, a group of experts that give us advice and feedback” (P21).

#### 3.5.2. Patient data and empirical evaluation

Many participants consider these semantic evaluation methods — reuse of existing concept sets and expert review — sufficient; others, however, hold that a value set for analytic use cannot be trusted without evaluating it or the phenotype or algorithm containing it through some form empirical review of clinical data:

> Chart Review. Some internal checking of codes against expected lab results, vital measurements, patient histories, etc. Ex. Diabetes codes should associate with histories of certain blood glucose measurements or A1C. (P28)

> First conduct discussion with clinical experts; Second, evaluate coverage of clinical concept in a data set; Third, perform random chart review to help detect if presence of code indicates disease. (P16)

> Lexical search, semantic exploration (navigate OHDSI vocab), empirical assessment thru characterization, and clinical expert review. (P05)

Every one of our 36 survey respondents reported that their *studies* use patient data. Nevertheless, of the 32 who answered the open-ended validation question, only nine indicated using patient data during value set development and validation. Choice of evaluation methods can be guided by clinical nuances of the research question or how the value set will be used. According to one survey respondent:

> Depends on the purpose and whether we are aiming for sensitivity or specificity. It may be chart review, or comparison with other **value set**s. (P34)

To unpack that statement a bit, a highly *sensitive* value set might be appropriate for instance for selecting patients to be screened for some condition where the goal is to capture as many patients as possible. When a sensitive value set is needed, comparison with existing value sets could help to make sure that appropriate codes are not missed. A highly *specific* value set may be suitable when recruiting patients for a clinical trial or when constructing the main cohort for an observational study, where false positives are costly. In this case, chart review of a sample of identified patients and value set/phenotype modifications will help.

Evaluating the accuracy of a sensitive value set requires a thorough semantic exploration to identify all codes that could indicate the condition of interest, while evaluating the accuracy of a specific value set should involve empirical examination of matching patient records to prevent false positives. (Software tools are available particularly to help value set developers discover codes related to the ones they start with: Term Sets [17] and PHenotype Observed Entity Baseline Endorsements (PHOEBE) [57]. PHOEBE functionality has recently been added to OHDSI’s ATLAS concept set editor.)

Table 5 lists general contexts in which value sets are used and relates them to the three literature categories listed above, to value set repositories that support them, and to the three conceptual dichotomies described in Sections 3.3, 3.4, 3.5: permissible/analytic, prescriptive/descriptive, and semantic/empirical.

### 3.6. A taxonomy of reasons for value sets to differ

While repositories make it possible to share and reuse value sets, clutter and redundancy can present serious challenges. For instance, a search for COPD (chronic obstructive pulmonary disease) on ATLAS (https://atlas-demo.ohdsi.org/#/conceptsets) gives 56 results. While many of these are usefully distinguished by their titles (e.g., Stage III-IV COPD or Concomitant COPD), many are not. In this section we break down the reasons that value sets for (ostensibly) the same clinical concept may differ into three categories: semantic, empirical, and due to error. These are summarized in Table 6.

**Table 6.** Reasons for value sets with same name to differ in definition and composition.

| VALID SEMANTIC | VALID EMPIRICAL | ERRONEOUS AND ARBITRARY |
| --- | --- | --- |
| Clinical meaning or nuance | Population characteristics | Codes mistakenly left out |
| Study requirements |  | Codes mistakenly included |
| Terminologies and cross-terminology mappings | Regional, institutional, or clinical specialization coding practices | Codes included or not based on faulty or idiosyncratic reference standards |
| Use of vocabularies lacking granularity for clinical concepts or requiring post-coordination | Institutional workflow |  |
| Algorithmic context (use of other domains or correcting for false positives or negatives at the phenotype level) |  |  |
|  | Arbitrary inclusion thresholds |  |

#### 3.6.1. Semantic reasons for value sets to differ

Two value sets may have the same name and refer to ostensibly the same condition or event but, on closer inspection, may differ in their meaning or how they are meant to be used.

##### Clinical meaning or nuance

Researchers may differ in their understanding of a clinical concept, or, for instance, a diabetes value set for a cardiology study may require a different set of codes than an endocrinology study.

##### Study requirements

Different value sets may share the same clinical meaning, but one study may need a more specific value set, another a more sensitive one.

##### Vocabulary issues

Value sets for the same phenomenon will, of course, differ if they use different vocabularies (e.g., ICD10CM or SNOMED-CT for clinical findings; NDC, RxNorm, or ATC for drugs; CPT, HCPCS, or ICD10PCS for procedures.) There may be reasons to translate codes across vocabularies. (E.g., CDMs like OMOP may require translation or harmonization of codes in patient records to agreed-upon vocabularies.) Different strategies may be applied when using vocabularies that either lack granularity to express the concept of interest or use post-coordination to express it.

##### Algorithmic context

Different strategies for identifying a clinical phenomenon can lead to differences in value set composition. (This not strictly a semantic issue but a fact of value set use.)

- A value set designed to target a diagnostic condition might use evidence from other domains of clinical data, e.g., drugs, procedures, lab tests. (It is recognized in the literature that phenotypes benefit from the use of multi-modal, multi-domain data [58].)
- A value set may be designed in the knowledge that it will produce false positives or negatives if these will be corrected by logic or other value sets at the phenotype.

#### 3.6.2. Empirical reasons for value sets to differ

Differences in the datasets being analyzed may affect the codes used to represent some conditions.

##### Population characteristics

E.g., codes may differ for studies of children less than 10 years of age versus geriatric populations; a study of neuropathy in orthopedic surgery patients would not need to include codes for diabetic neuropathy in the pediatric population.

##### Regional, institutional, or clinical specialization coding practices

A single meaning may be expressed using different codes in different places. (This possibility was mentioned by our participants and seems to be a relatively common belief, but we have not encountered specific examples.)

##### Institutional workflow at data source

Certain conditions or observations may not be captured in some clinical settings requiring recourse to indirect ways of identifying them in EHRs.

##### Arbitrary inclusion thresholds

Some codes may give rise to false positives when included and false negatives when left out. If researchers are not able to resolve this kind of problem at the phenotype algorithm level, they will need to make judgment calls depending on whether they think the false negatives or false positives caused by a given code’s presence or absence will more adversely affect the study’s results. Differences in judgment do not mean one decision is right and another wrong, but, unless the judgment call is justified with specific reasoning, value sets for a given phenomenon can differ without giving potential re-users any basis for choosing between them.

#### 3.6.3. Errors

Crafting accurate value sets is hard and mistakes are not uncommon. Discrepancies between value sets provide an opportunity to discover mistakes that might otherwise be overlooked.

##### Codes mistakenly left out or mistakenly included

When value sets are missing codes they should include, they can cause false negatives in patient or event selection; codes included in error can introduce false positives in selection. Without a reference (or gold) standard to test results (of a value set or its containing cohort algorithm) against a sample of records already reliably classified as exhibiting or not exhibiting the clinical phenomenon of interest, false positives and negatives in selection results may entirely escape detection.

##### Codes included or not based on faulty or idiosyncratic reference standards

Reference standards themselves can suffer from error. Decisions by a chart reviewer on which patients match a phenotype or cohort definition can be affected by differences in understanding that are not quite matters of clinical judgment or study needs but differences in chart reading practice, differences in the chart reviewers’ interpretation of study needs, or chart reviewer error. But if the error or discrepancy affects the standard, a value set or its containing phenotype may show perfect sensitivity (low false negative rate) and specificity (low false positive rate) while differing from a value set based on another gold standard.

Errors can lead to bias in results, whose magnitude and direction are not predictable, but legitimate differences in value sets can be recognized if their reasons are known. Value set analysis and authoring software can be better designed to help re-users understand these differences, giving them a basis for deciding between existing value sets or selecting the elements from each most appropriate for their own use case.

### 3.7. Conclusion: Leveraging and mitigating redundancy in value set repositories

Value set reuse is frequently championed as a response to persistent concerns about value set quality: not only should researchers make use of expertly designed value sets, value set repositories should facilitate incremental refinement; over time the quality of a shared value set should improve as more researchers put it to use, evaluate its accuracy, and contribute their changes back to the repository.

> [Reusable value sets] would be helpful [so that] I don’t have to do this on my own every time…[B]ecause it has been created by a collaborative team that’s known for creating value sets, I would know that, "Oh, this has been extracted or they got it from a paper that has been vetted and validated and you know it’s a legit paper." I would use that. (P09, interview)

Rather than incremental improvement of existing value sets or indications of a value set’s having been vetted and validated, what we see in repositories is proliferation and clutter: new value sets that may or may not have been vetted in any way and junk concept sets, created for some reason but never finished. We have found general agreement in our data that the presence of many alternative value sets for a given condition often leads value set developers to ignore all of them and start from scratch, as there is generally no easy way to tell which will be more appropriate for the researcher’s needs. And if they share their value set back to the repository (as they must on analysis platforms like ATLAS or N3C), they further compound the problem, especially if they neglect to document the new value set’s intention and provenance.

There is a tension regarding how many value sets should exist for a given clinical condition. On the one hand, the principle of reproducibility of research and fungibility of research results—whether results from different studies may be pooled—argues for re-use of value sets. On the other hand, tight coherence with the research question — “fitness for use” — argues for customizing a unique value set to fit the research intent. Given this tension, it is no surprise that respondents expressed a variety of beliefs on each side of this dialectic.

If, as a field, we hope to increase reuse and refinement to decrease redundant value set creation, we must be able to understand when an additional value set for a target condition may be needed or not. The taxonomy in Section 3.6 may help in reconciling differences when multiple value sets are being reviewed or considered for reuse: if the analyst can identify a valid reason for a difference, this may give them insight to inform choices for their own use case or may help them determine where errors lie, increasing or decreasing their confidence in specific value sets or codes.

When a new researcher creates their own value set from scratch rather than leveraging the work of those who have tread the same ground, however, this should not be seen as laziness or as a problem to be addressed by exhortations to reuse existing value sets. Rather, the fault should be ascribed to the resources available to them: they should be given software and metadata to make the review and comparison of existing value set easier than creation from scratch. Practical application of the taxonomy and other ideas presented in this paper will require new software designed to implement these ideas and better guide value set developers through the process. Toward that end, we offer the following recommendations.

#### 3.7.1. Advanced, automated comparison tools

In our professional experience we have seen instances where trust in what were considered authoritative value sets broke down when comparing them to other value sets. One participant, P16, performed an automated comparison of many alternative value sets for depression, using the differences and similarities to create a trustable value set without having to trust any of the input value sets individually.

Comparison functionality should be a central feature of value set repositories and authoring platforms, allowing users to take advantage of existing value sets rather than burdening them with having to manually sift through ostensibly redundant value sets. In the last couple years, tools for comparing value sets have begun to appear in ATLAS, the N3C Concept Set Browser, and VS-Hub [59]. VS-Hub explicitly nudges the user to compare related value sets and highlights the selected value sets’ similarities and differences throughout authoring and review.

#### 3.7.2. Detailed metadata collection and use

Existing tools vary in their collection of metadata through the authoring process, but however much metadata they collect, it is at the value set level; it could be enormously helpful to collect metadata to capture value set developers’ reasoning for including or rejecting specific codes. (FHIR and N3C accommodate relatively extensive set of metadata fields; VSAC somewhat less; and OHDSI/ATLAS hardly any at all. N3C, at SG’s suggestion, does request reasoning when adding codes to a value set, but this feature has not yet been developed to the point of being useful — nothing is currently done with users’ input.) A combination of automated process data capture and timely, minimally obtrusive user prompts could provide code-level metadata that could be displayed as future value set authors consider whether to include a code or not. An automated capture process could, for instance, record the source of included codes: if found in an existing value set, record a reference to that value set; if found through vocabulary text search and/or navigation of vocabulary hierarchy, record the steps leading to the included code. User prompts could try, for instance, to capture whether patient counts or any kind of chart review or gold standard had been used in decisions to include or reject specific codes.

#### 3.7.3. Expert or automated curation

Terminology experts on the N3C infrastructure staff have developed “N3C Recommended” value sets for commonly studied topics (conditions, medications, medication classes, measurements, procedures.) VS-Hub was specifically designed to facilitate that endeavor. VS-Hub, so far, has only attempted to make it easier to cull the best out of available value sets for a given condition or event, it has not attempted to force users to review relevant value sets and either improve one of those or make sure that a new one is genuinely needed. If we, as a field, hope to see value set repositories increase rather than decrease in quality and usefulness over time and widening use, strong curation will be necessary to exclude redundant, unfinished, or otherwise low-quality value sets. Such curation could be done by humans, software, or both.

The requirements and recommendations in prior literature have not been sufficient to guide the design of software that could make effective leveraging of shared value sets a reality. However, the conceptual framework, real-world experience, and deep, detailed account of the challenges to reuse presented here make up that deficit and provide a high-level requirements roadmap for improved code-set creation tools.

## Data Availability

Survey data have been anonymized and made available at Gold S. Value sets and the problem of redundancy in value set repositories. Survey data. OSF. 2024. doi:10.17605/OSF.IO/ABTJU
Interview data cannot be anonymized and are not included to protect participant privacy.

https://osf.io/abtju/

## CRediT authorship contribution statement

**Sigfried Gold**: Conceptualization, Methodology, Formal analysis, Investigation, Data Curation, Writing - Original Draft, Writing - Review & Editing, Visualization; **Harold P. Lehmann**: Writing - Review & Editing, Supervision; **Lisa M. Schilling**: Writing - Review & Editing, Supervision; **Wayne G. Lutters**: Methodology, Writing - Original Draft, Writing - Review & Editing, Supervision, Project administration

## Acknowledgments

Richard Williams, Jessica Ancker, Christopher Chute, Davera Gabriel, Harold Solbrig, Jeff Brown, Laura Wiley, Luke Rasmussen, Rachel Richesson, Allen Flynn, Meredith Zozus, David Gotz, Erica Voss, Christian Reich, Kristin Kosta, Shelley Rusincovitch, Niklas Elmqvist, Leilani Battle, Joel Chan, Amanda Lazar.

